# Dietary Restraint Moderates the Relationship between Postprandial Distress Syndrome and Binge Eating in Adolescents

**DOI:** 10.1101/2025.09.03.25335031

**Authors:** Laura Rubino, Elizabeth W. Lampe, Sophie R. Abber, Stephanie M. Manasse

**Affiliations:** Department of Psychological and Brain Sciences, Drexel University; Center for Technology & Behavioral Health, Department of Quantitative Biomedical Data Science, Geisel School of Medicine, Dartmouth College; Department of Psychology, Florida State University; Department of Psychiatry, University of California, San Diego; Center for Healthcare Delivery Science, Nemours Children’s Health

**Keywords:** Binge Eating, Gastrointestinal Symptoms, Dietary Restraint, Overweight, Obesity, Eating Disorders

## Abstract

**Objective:** The current study investigated whether dietary restraint moderates the relationship between Postprandial Distress Syndrome (PDS) symptoms (e.g., postprandial fullness, early satiety, and nausea) and Binge Eating (BE) in adolescents with overweight and obesity. BE is associated with PDS symptoms. Individuals often self-initiate restrictive diets that limit food variety in attempt to manage PDS symptoms. However, dietary restraint is thought to increase frequency of BE. Thus, individuals who attempt to manage PDS symptoms through dietary restraint may engage in more BE. Investigating the relationship between dietary restraint and PDS in relation to BE will increase our understanding of risk factors for BE during adolescence—a period associated with heightened onset of disordered eating and dieting attempts.

**Method:** Adolescents (*N*=60) with higher weight (*M* Body Mass Index (BMI) percentile = 97.72 [*SD* = 2.61], *M* age = 15.37 years [SD = 1.34], 63.3% female) completed measures of PDS symptoms, dietary restraint, and BE. Generalized linear regression examined whether dietary restraint moderated the relationship between PDS symptoms and BE frequency.

**Results:** The interaction effect was significant, such that greater PDS symptoms were associated with increased BE frequency when dietary restraint was average or higher than average.

**Discussion:** Results suggest that the relationship between PDS and BE symptoms depends on level of dietary restraint. Future work should investigate if this relationship holds for other GI syndromes and investigate the differential impact of dietary restraint versus restriction on PDS and BE.

## Introduction

Binge eating (BE) is a transdiagnostic eating disorder symptom characterized by consuming a large amount of food in a short period of time and is most commonly associated with Binge Eating Disorder (BED; American Psychiatric Association, 2013). BE is associated with greater symptoms of Functional Dyspepsia (FD; Hanel et al., 2021; Santonicola, 2012) and 53.6% of individuals with BED report FD symptoms (Félix-Téllez et al., 2025). Specifically, BED has been associated with a 52% increased likelihood of postprandial distress syndrome (PDS) symptoms, a subtype of FD characterized by postprandial fullness, early satiety, and nausea (Drossman & Hasler, 2016; Félix-Téllez et al., 2025). In one sample of adults with BED, 38.6% endorsed early satiety and 36.2% endorsed postprandial fullness (Félix-Téllez et al., 2025). This association appears to be independent of obesity’s effects on GI symptoms. Among adults with obesity, those who endorsed BE reported greater PDS symptoms (Crowell et al., 1994; Santonicola et al., 2013). Understanding for whom PDS symptoms might be associated with BE will increase our understanding of who is at risk for BE and inform prevention interventions.

BE is defined by a specific pattern of eating (e.g., eating an excessive amount of food in a short period of time). PDS symptoms typically occur after eating episodes and the overeating associated with BE could exacerbate PDS symptoms (Murray et al., 2021; Santonicola, 2012). In response to PDS and FD symptoms, individuals often attempt self-guided diet manipulations such as caloric restriction or avoiding foods believed to trigger GI symptoms (Atkins et al., 2023; Pucci & Forney, 2022; Reed-Knight et al., 2016; van Tilburg & Felix, 2013). This dietary restraint—the cognitive effort of restricting calories or food intake, regardless of the behavioral outcome of the restraint—has been associated with higher rates of BE. (Andrés & Saldaña, 2014; Schaumberg et al., 2016). The increased cognitive focus on food and overreliance of cognitive control to dictate eating versus physiological cues, can increase the risk of BE and overeating (Herman & Polivy, 1980). Thus, individuals with PDS may be at increased risk for BE, particularly when they engage in dietary restraint intended to reduce GI symptoms. As patients often attempt self-initiate diets or specific patterns of eating to manage PDS symptoms, it is important to clarify whether dietary restraint increases the risk of BE among those with PDS symptoms (Duboc et al., 2020; Weeks et al., 2023). This relationship may be particularly relevant during adolescence—a heightened period of risk for onset of BE and often a period when individuals attempt dieting for the first time (Marzilli et al., 2018; Schaumberg et al., 2016).

To further our understanding of potential factors that might increase risk BE among adolescents experiencing PDS symptoms, the current study aimed to investigate the relationship between PDS symptoms, dietary restraint, and BE in adolescents. We hypothesized that dietary restraint would moderate the relationship between PDS symptoms and BE, such that PDS symptoms would be more strongly associated with BE episodes at greater levels of dietary restraint.

## Methods

### Participants and procedures

Participants (*N*=60) were adolescents with higher weight (*M* BMI percentile = 97.72 (*SD* = 2.61)) enrolled in a pilot study evaluating a lifestyle modification program (K23DK124514). Eligible participants were between the ages of 14 and 18, had a BMI >= 85^th^ percentile for sex and age, living at home with a parent/guardian, had at least one parent willing to participate in the study, lived in the United States, and were willing to complete study procedures. Exclusion criteria included acute suicide risk, inability to engage in physical activity (defined as walking two city blocks without stopping), diabetes, history of bariatric surgery, current medical or psychiatric condition that could interfere with intervention participation, pregnant or planning to become pregnant, weight loss of greater than 10% in the previous 6 months, engagement in compensatory vomiting after BE or more than 12 of any compensatory behaviors in the previous 3 months, plans to start another weight loss treatment or psychotherapy for BE, and currently taking weight-loss medications. Eligible participants completed a virtual visit in which informed assent/consent was obtained, height and weight was measured, and the Eating Disorder Examination (EDE) was administered by a trained research assistant. After completion of the virtual visit, participants completed a battery of self-report measurements at home. All procedures were approved by the Drexel University Institutional Review Board (Protocol Number 2005007834A002).

### Measures

#### Postprandial Distress Syndrome

Participants completed the Gastrointestinal Symptoms Questionnaire (GISQ; Bovenschen et al., 2006), a 32-item questionnaire assessing GI symptom severity over the past four weeks. Each question assessed a different GI symptom (e.g., heartburn, regurgitation) on a 7-point Likert-type scale (1 “none” to 7 “unbearable”). We created a composite PDS score by summing the nausea, vomiting, loss of appetite, and postprandial fullness items (α=.65). Scores could range from 4 to 28, with higher scores indicating greater symptom severity.

#### Binge Eating and Dietary Restraint

The Eating Disorders Examination 17.0 (EDE; Thomas et al., 2014) was used to assess BE frequency and dietary restraint over the previous 12 weeks. BE was defined as the total number of objective and subjective binge episodes experienced over the past 12 weeks. To measure restraint we used the EDE Restraint subscale, which is the average of five items: conscious attempts to reduce caloric intake, avoidance of eating, avoidance of certain foods, following dietary rules, and desiring an empty stomach. Scores can range from 0.0 to 6.0 with higher scores indicating greater restraint (α=0.65).

### Statistical Analysis

We used a Compound Poisson Generalized Linear Model with (CPGLM) to analyze the count data in this study. The CPGLM model was appropriate due to the over-dispersion and zero-inflation observed in frequency of BE, the outcome variable. The PDS composite score was the independent variable, dietary restraint the moderator, and we covaried for birth sex and age. We did not covary for BMI, given the restricted range observed in the sample, with inclusion requiring > 85^th^ percentile and most participants were in the 99^th^ percentile. We used the cplm package in R (Zhang, 2013). We conducted simple slopes analyses. Levels of the moderator (i.e., dietary restraint) were categorized as low (i.e., more than one standard deviation (SD) below the mean), average (i.e., within 1 SD of the mean), and high (i.e. greater than one SD above the mean).

## Results

### Participant Characteristics

Participants (N = 60) were 14-18 years old, (*M* = 15.37 years, *SD* = 1.34 years). Thirty-eight (63.3%) were assigned female at birth and 37 (61.7%) identified as women. Of the 60 participants, 63.3% identified as White, 16.7% Black or African American, 5% American Indian/Alaska Native, 1.7% Asian, 10% multiracial, and 3.3% were unknown or chose not to report race. Additionally, 8.3% identified as Hispanic/Latine. Of the 60 participants, 21 (35%) reported experiencing at least one subjective or objective BE episode over the past three months (*M* = 11.57, *SD* = 24.83, range = 0 - 110).

### Primary Aim

The main effect between PDS and BE was non-significant. The main effect between dietary restraint and BE was also non-significant. The interaction between PDS and dietary restraint was significant indicating that the effect of PDS symptoms on BE varied depending on the level of dietary restraint (Table 1). Specifically, the relationship between PDS and BE was strongest for those with the highest levels of dietary restraint. For individuals in the highest tertile of dietary restraint, every 2-point increase in PDS symptoms was associated with 1 more BE episode over the past 3 months (Figure 1). In the lowest tertile of dietary restraint, PDS symptoms were not significantly correlated to BE episodes (b = 0.11, SE = 0.10, t = 1.17, *p* =.25). In the both the middle tertile (b = 0.31, SE = 0.10, t = 3.26, *p*=.002), and highest tertile (b= 0.52, SE = 0.16, t = 3.15, *p*=.003), PDS symptoms significantly predicted BE episodes.

**Table 1.**
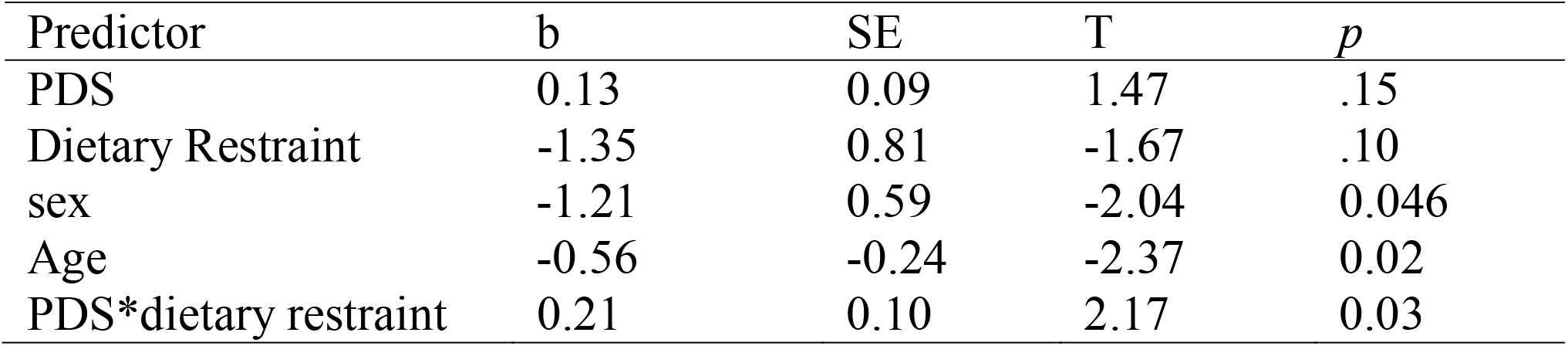
Compound Poisson Generalized Linear Model (CPGLM) results with postprandial distress syndrome (PDS) composite as the independent variable, dietary restraint as the moderator, and binge eating episodes as the outcome. Sex and age were covariates.

| Predictor | b | SE | T | *p* |
| --- | --- | --- | --- | --- |
| PDS | 0.13 | 0.09 | 1.47 | .15 |
| Dietary Restraint | -1.35 | 0.81 | -1.67 | .10 |
| sex | -1.21 | 0.59 | -2.04 | 0.046 |
| Age | -0.56 | -0.24 | -2.37 | 0.02 |
| PDS*dietary restraint | 0.21 | 0.10 | 2.17 | 0.03 |

**Figure 1.**
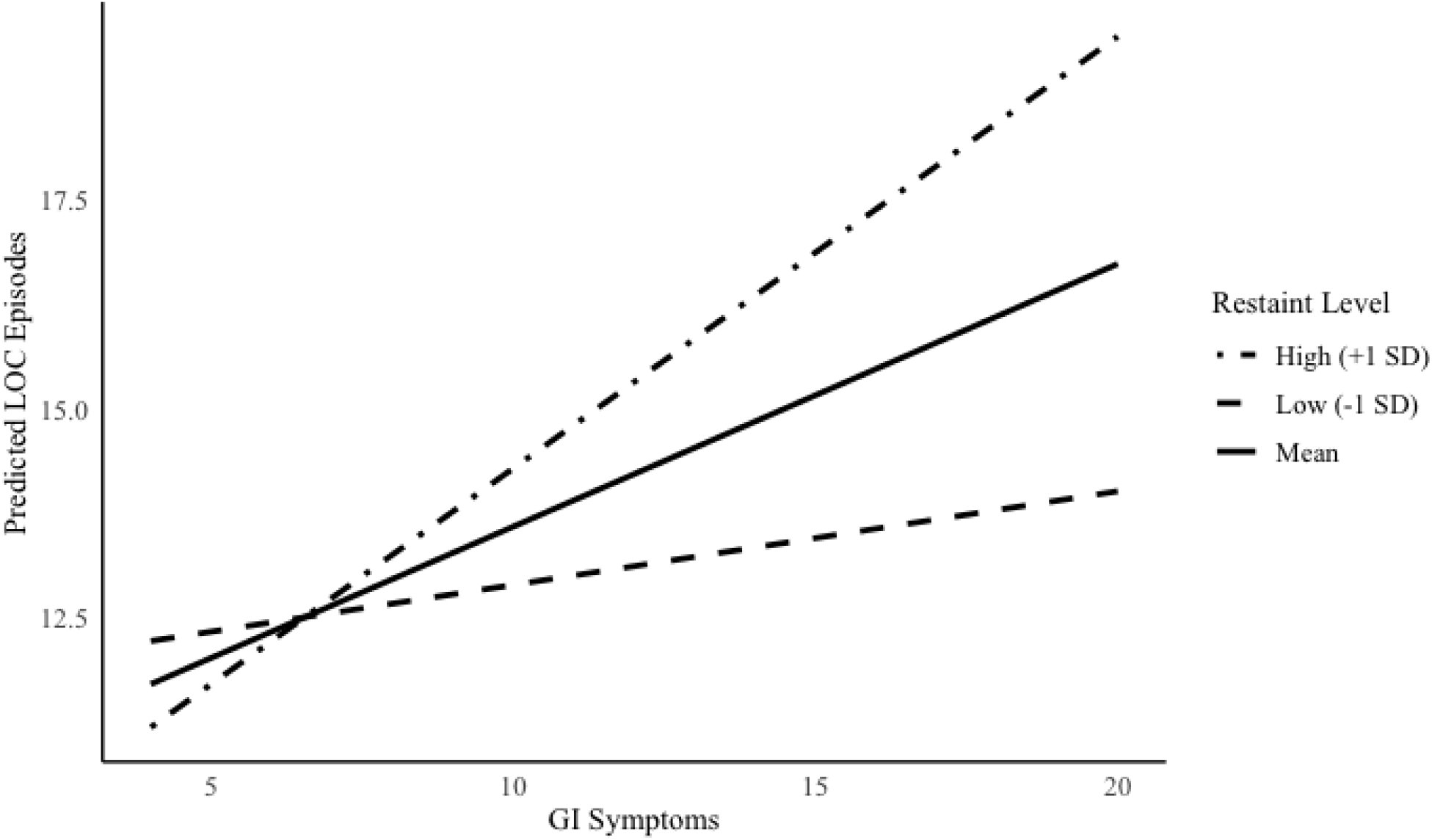
Simple Slopes of GI Symptoms at Different Levels of Restraint.

### Post-hoc Analysis

Data were cross-sectional, making it difficult to determine the direction of the relationship. To address this, we tested the reciprocal relationship with PDS composite as the dependent variable, BE episodes as the independent variable, dietary restraint as the moderator to examine whether the effect might operate in the opposite direction from our initial moderation model. We conducted a second CPGLM model. We covaried for birth sex and age. Main effects were non-significant with BE (b = 0.003, SE = 0.003, t = 1.01, *p* =.32) and dietary restraint (b = 0.04, SE = 0.06, t = 0.76, *p* =.45) not predicting PDS. The interaction effect was also non-significant (b = −0.0005, SE = 0.002, t = −0.21, *p* =.83).

## Discussion

The current study investigated a potential moderator between PDS symptoms and BE with adolescents with overweight or obesity, increasing understanding about who with PDS might be at increased risk for BE. Consistent with our hypothesis, the current study found there was a significant interaction between PDS and dietary restraint. The relationship between PDS and BE depended on dietary restraint, such that there was a strong positive relationship between PDS and BE when dietary restraint was average or higher than average, but not when dietary restraint was lower than average.

Due to the tendency to respond to PDS symptoms with diet manipulations, some adolescents who experience PDS symptoms may engage in dietary restraint. Current results suggest that among adolescents experiencing PDS symptoms, high dietary restraint increases BE risk. Dietary restraint heightens cognitive focus on and preoccupation with food which is cognitively taxing. It is theorized this cognitive tax diminishes self-control over time, contributing to later BE episodes (Loth et al., 2016). Further, individuals with PDS may engage in hedonic restraint—avoiding foods that they find pleasurable. Research theorizes that hedonic restriction can lead to binge eating (Lowe et al., 2016; Lowe & Butryn, 2007; Witt & Lowe, 2014). Interventions for PDS should therefore try to minimize unnecessary diet manipulations. For instance, when foods are not avoided due to a true allergy or structural GI condition, treatment could include psychoeducation that eating feared foods can help alleviate GI symptoms (Biesiekierski et al., 2022; Keefer et al., 2022; Murray et al., 2021) and reduce BE (Linardon et al., 2023).

Strengths of the current study include the use of a community adolescent sample and gold-standard measures of both BE and restraint. Limitations of the current study include low symptom endorsement of PDS symptoms, low endorsement of BE in the sample, and use of a non-validated measure of PDS symptoms. Both the PDS composite and EDE Restraint had acceptable, albeit somewhat low, internal consistency. Further, this study was cross-sectional and although we hypothesized that it was dietary restraint that moderated the relationship, directionality cannot be confirmed.

Future research should replicate these findings in a sample with greater symptom endorsement and use a measure specifically designed to capture PDS symptoms. Replicating these findings in a larger sample and longitudinal design will help confirm the strength and directionality of the relationship. Future work should also tease apart the impact of dietary restraint versus dietary restriction on the relationship between BE and PDS symptoms and work to understand whether restriction plays a role above and beyond restraint in the relationship between BE and PDS. It will also be important to learn who with PDS symptoms is at greater risk for higher dietary restraint and if this relationship exists for other GI syndromes as well. Overall, the current study provides preliminary evidence that adolescents with more severe PDS symptoms are at greater risk for BE when they also engage in high levels of dietary restraint.

## Data Availability

All data produced in the present study are available upon reasonable request to the authors

## References

American Psychiatric Association. (2013). Diagnostic and Statistical Manual of Mental Disorders (Fifth Edition). American Psychiatric Association. 10.1176/appi.books.9780890425596

Andrés, A., & Saldaña, C. (2014). Body dissatisfaction and dietary restraint influence binge eating behavior. Nutrition Research, 34(11), 944–950. 10.1016/j.nutres.2014.09.003

Atkins, M., Zar-Kessler, C., Madva, E. N., Staller, K., Eddy, K. T., Thomas, J. J., Kuo, B., & Burton Murray, H. (2023). History of trying exclusion diets and association with avoidant/restrictive food intake disorder in neurogastroenterology patients: A retrospective chart review. Neurogastroenterology and Motility, 35(3), e14513. 10.1111/nmo.14513

Biesiekierski, J. R., Manning, L. P., Murray, H. B., Vlaeyen, J. W. S., Ljótsson, B., & Van Oudenhove, L. (2022). Review article: Exclude or expose? The paradox of conceptually opposite treatments for irritable bowel syndrome. Alimentary Pharmacology & Therapeutics, 56(4), 592–605. 10.1111/apt.17111

Bovenschen, H. J., Janssen, M. J. R., van Oijen, M. G. H., Laheij, R. J. F., van Rossum, L. G. M., & Jansen, J. B. M. J. (2006). Evaluation of a gastrointestinal symptoms questionnaire. Digestive Diseases and Sciences, 51(9), 1509–1515. 10.1007/s10620-006-9120-6

Crowell, M. D., Cheskin, L. J., & Musial, F. (1994). Prevalence of gastrointestinal symptoms in obese and normal weight binge eaters. The American Journal of Gastroenterology, 89(3), 387–391.

Drossman, D. A., & Hasler, W. L. (2016). Rome IV—functional GI disorders: Disorders of gut-brain interaction. Gastroenterology, 150(6), 1257–1261. 10.1053/j.gastro.2016.03.035

Duboc, H., Latrache, S., Nebunu, N., & Coffin, B. (2020). The Role of Diet in Functional Dyspepsia Management. Frontiers in Psychiatry, 11, 23. 10.3389/fpsyt.2020.00023

Félix-Téllez, F. A., Cruz-Salgado, A. X., Remes-Troche, J. M., Flores-Rendon, Á.R., Ordaz-Álvarez, H. R., Velasco, J. A. V.-R., Flores-Lizárraga, M. A. O., Soto-González, J. I., & Abizaid-Herrera, N. S. (2025). Association Between Functional Dyspepsia and Binge Eating Disorder: A Frequent, Often Overlooked Overlap Clinical Presentation. Journal of Neurogastroenterology and Motility, 31(1), 95–101. 10.5056/jnm24070

Hanel, V., Schalla, M. A., & Stengel, A. (2021). Irritable bowel syndrome and functional dyspepsia in patients with eating disorders □ a systematic review. European Eating Disorders Review, 29(5), 692–719. 10.1002/erv.2847

Herman, C. P., & Polivy, J. (1980). Restrained Eating. In A. J. Standard (Ed.), Obesity, 208–225. Philadelphia: W.B. Saunders

Keefer, L., Ballou, S. K., Drossman, D. A., Ringstrom, G., Elsenbruch, S., & Ljótsson, B. (2022). A Rome Working Team Report on Brain-Gut Behavior Therapies for Disorders of Gut-Brain Interaction. Gastroenterology, 162(1), 300–315. 10.1053/j.gastro.2021.09.015

Linardon, J., Messer, M., Shatte, A., Skvarc, D., Rosato, J., Rathgen, A., & Fuller-Tyszkiewicz, M. (2023). Targeting dietary restraint to reduce binge eating: A randomised controlled trial of a blended internet- and smartphone app-based intervention. Psychological Medicine, 53(4), 1277–1287. 10.1017/S0033291721002786

Loth, K. A., Goldschmidt, A. B., Wonderlich, S. A., Lavender, J. M., Neumar □ Sztainer, D., & Vohs, K. D. (2016). Could the resource depletion model of self □ control help the field to better understand momentary processes that lead to binge eating? International Journal of Eating Disorders, 49(11), 998–1001. 10.1002/eat.22641

Lowe, M. R., Arigo, D., Butryn, M. L., Gilbert, J. R., Sarwer, D., & Stice, E. (2016). Hedonic hunger prospectively predicts onset and maintenance of loss of control eating among college women. Health Psychology, 35(3), 238–244. 10.1037/hea0000291

Lowe, M. R., & Butryn, M. L. (2007). Hedonic hunger: A new dimension of appetite? Physiology & Behavior, 91(4), 432–439. 10.1016/j.physbeh.2007.04.006

Marzilli, E., Cerniglia, L., & Cimino, S. (2018). A narrative review of binge eating disorder in adolescence: Prevalence, impact, and psychological treatment strategies. Adolescent Health, Medicine and Therapeutics, Volume 9, 17–30. 10.2147/AHMT.S148050

Murray, H. B., Kuo, B., Eddy, K. T., Breithaupt, L., Becker, K. R., Dreier, M. J., Thomas, J. J., & Staller, K. (2021). Disorders of gut–brain interaction common among outpatients with eating disorders including avoidant/restrictive food intake disorder. International Journal of Eating Disorders, 54(6), 952–958. 10.1002/eat.23414

Pucci, G., & Forney, K. J. (2022). Associations among perceived taste and smell sensitivity, gastrointestinal symptoms, and restrictive eating in a community sample of adults. Eating Behaviors, 46, 101647. 10.1016/j.eatbeh.2022.101647

Reed-Knight, B., Squires, M., Chitkara, D. K., & van Tilburg, M. a. L. (2016). Adolescents with irritable bowel syndrome report increased eating-associated symptoms, changes in dietary composition, and altered eating behaviors: A pilot comparison study to healthy adolescents. Neurogastroenterology and Motility, 28(12), 1915–1920. 10.1111/nmo.12894

Santonicola, A. (2012). Prevalence of functional dyspepsia and its subgroups in patients with eating disorders. World Journal of Gastroenterology, 18(32), 4379. 10.3748/wjg.v18.i32.4379

Santonicola, A., Angrisani, L., Ciacci, C., & Iovino, P. (2013). Prevalence of Functional Gastrointestinal Disorders according to Rome III Criteria in Italian Morbidly Obese Patients. The Scientific World Journal, 2013(1), 532503. 10.1155/2013/532503

Schaumberg, K., Anderson, D. A., Anderson, L. M., Reilly, E. E., & Gorrell, S. (2016). Dietary restraint: What’s the harm? A review of the relationship between dietary restraint, weight trajectory and the development of eating pathology. Clinical Obesity, 6(2), 89–100. 10.1111/cob.12134

Thomas, J. J., Roberto, C. A., & Berg, K. C. (2014). The Eating Disorder Examination: A semi-structured interview for the assessment of the specific psychopathology of eating disorders. Advances in Eating Disorders, 2(2), 190–203. 10.1080/21662630.2013.840119

van Tilburg, M. A. L., & Felix, C. T. (2013). Diet and functional abdominal pain in children and adolescents. Journal of Pediatric Gastroenterology and Nutrition, 57(2), 141–148. 10.1097/MPG.0b013e31829ae5c5

Weeks, I., Abber, S. R., Thomas, J. J., Calabrese, S., Kuo, B., Staller, K., & Murray, H. B. (2023). The intersection of disorders of gut-grain interaction with avoidant/restrictive food intake disorder. Journal of Clinical Gastroenterology, 57(7), 651–662. 10.1097/MCG.0000000000001853

Witt, A. A., & Lowe, M. R. (2014). Hedonic hunger and binge eating among women with eating disorders. International Journal of Eating Disorders, 47(3), 273–280. 10.1002/eat.22171

